# Morning exercise and pre-breakfast metformin interact to reduce glycaemia in people with Type 2 Diabetes: a randomized crossover trial

**DOI:** 10.1101/2023.09.07.23295059

**Authors:** Brenda J. Pena Carrillo, Emily Cope, Sati Gurel, Andres Traslosheros, Amber Kenny, Nimesh Mody, Mirela Delibegovic, Sam Philip, Frank Thies, Dimitra Blana, Brendan M. Gabriel

**Author notes:** Corresponding author – Dr B.M. Gabriel, P: The Rowett Institute, University of Aberdeen, Ashgrove Rd W, Aberdeen AB25 2ZD, Aberdeen, UK. E.

## Abstract

Exercise is recommended in the treatment of Type 2 Diabetes and can improve insulin sensitivity [1]. However, previous evidence suggests that exercise at different times of the day in people with type 2 diabetes may have opposing outcomes on glycaemia [2]. Metformin is the most commonly prescribed initial pharmacological intervention in Type 2 Diabetes, and may alter adaptions to exercise [3]. It is unknown if there is an interaction between metformin and diurnal exercise outcomes. We aimed to investigate glycaemic outcomes of moderate intensity morning vs. evening exercise in people with type 2 diabetes being prescribed metformin monotherapy. We hypothesised that evening exercise would be more efficacious than morning exercise at lowering glycaemia.

In this study, nine male and nine females with type 2 diabetes undergoing metformin monotherapy (age 61±2 year, mean±SEM) completed a 16-week crossover trial including 2- week baseline recording, six weeks randomly assigned to a morning exercise (7-10am) or evening exercise (4-7pm), and a two-week wash-out period. Exercise arms consisted of 30 minutes of walking at 70% of estimated max-HR every other day. Glucose levels were measured with continuous glucose monitors and activity measured by wrist-worn monitors. Food-intake was recorded by 4-day food diaries during baseline, first and last 2 weeks of each exercise arm.

There was no difference in exercise intensity, total caloric intake, or total physical activity between morning and evening arms. Acute glucose area under the curve (AUC), was lower (*p*=0.02) after acute morning exercise (180.6±16.1 mmol/L) compared to baseline (210.3±18.0 mmol/L). Acute AUC glucose was significantly lower (*p*=0.01) in participants taking metformin before breakfast (152.5±10.59 mmol/L) compared with participants taking metformin after breakfast (227.2±27.51 mmol/L) only during the morning exercise arm. During weeks 5-6 of the exercise protocol, AUC glucose was significantly lower (p=0.04) for participants taking metformin before breakfast (168.8±5.6), rather than after breakfast (224.5±21.2) only during morning exercise.

Our data reveal morning moderate exercise acutely lowers glucose levels in people with type 2 diabetes being prescribed metformin. This difference appears to be driven by individuals that consumed metformin prior to breakfast rather than after breakfast. This beneficial effect upon glucose levels of combined morning exercise and pre-breakfast metformin persisted through the final two weeks of the trial. Our findings suggest that morning moderate intensity exercise combined with pre-breakfast metformin intake may benefit the management of glycaemia in people with type 2 diabetes.

**Research in Context:** *What is already known about this subject?:*

- Exercise at different times of the day in people with Type 2 Diabetes has opposite outcomes on glycaemia.
- Metformin interferes with the glucose-lowering effect of acute exercise.
- It is unknown if there is an interaction between metformin intake timing and diurnal exercise outcomes.

*What is the key question?:*

- Is it possible to optimise timing of concomitant metformin and exercise in people with Type 2 Diabetes?

*What are the new findings?:*

- Morning moderate exercise acutely lowers glucose levels in people with Type 2 Diabetes being prescribed metformin.
- This difference appears to be driven by individuals that consumed metformin prior to breakfast rather than after breakfast.
- Morning exercise combined with pre-breakfast metformin persistently reduced glucose compared to morning exercise combined with post-breakfast metformin through the final week (week 6) of the intervention.

*How might this impact on clinical practice in the foreseeable future?:*

- Our study suggests it may be possible to make simple changes to the time that people with Type 2 Diabetes take metformin and perform exercise to improve their blood glucose.

## 1. Introduction

Type 2 Diabetes Mellitus is a growing, global health challenge [4]. Exercise is often recommended by front-line clinicians as a potent therapeutic treatment in people with type 2 diabetes which can improve insulin sensitivity [1]. However, little is known regarding the optimal time of day to perform exercise for people with Type 2 Diabetes. Previous evidence suggests that morning high intensity interval training over two weeks increases blood glucose levels in people with type 2 diabetes [2], drawing parallels to the dawn phenomenon. Conversely, evening exercise reduced glycaemia on the day of exercise in people with type 2 diabetes [2]. These apparently divergent outcomes of morning vs. evening exercise highlight that it is imperative to optimise the timing of exercise for people with type 2 diabetes.

Several other studies have assessed the time-of-day effect of exercise upon glycaemic or insulin-related outcomes. For example, cross-sectional data in 775 participants demonstrates that moderate-to-vigorous exercise in the afternoon or evening was associated with a greater reduction of insulin resistance compared with an evenly distributed daily time-pattern of exercise [5]. Further associative evidence [6], demonstrates that in 2,416 participants that moderate-to-vigorous physical activity performed in the afternoon is associated with improvements in glycaemic control in people with diabetes. In a randomised trial, men who were overweight or obese demonstrated improvements in glycaemic control after evening exercise, while a group who exercised in the morning saw no improvements in glycaemic control [7]. In another randomised trial assessing people with compromised metabolism, a group that performed exercise training in the evening experienced superior beneficial improvements in fasting plasma glucose levels compared to a group that performed exercise training in the morning [8]. Skeletal muscle is key in the metabolic response to exercise [1] and it has been demonstrated that people with type 2 diabetes have an intrinsically altered circadian rhythm in skeletal muscle, both at the level of the molecular clock, and of mitochondrial function [9]. Therefore, it is not clear whether the divergent time-of-day effect of exercise on glycaemic regulation is due to an intrinsic metabolic disruption, or whether this phenomenon will persist in people who are metabolically healthy. Indeed, glycaemic regulation does not appear to be different in metabolically healthy participants between a single bout of morning or afternoon exercise [10].

Metformin is the most commonly prescribed initial pharmacological intervention in type 2 diabetes and is known to alter skeletal muscle mitochondrial adaptions to exercise [3]. Further, it has been suggested that metformin may interfere with the glucose-lowering effect of acute exercise [11, 12]. However, it is unclear whether there is an interaction between metformin and exercise at different times of the day. Optimising exercise timing in people with type 2 diabetes is critically important, but also is to understand any interactions with widely prescribed pharmacological treatments. Furthermore, previous trials [2, 13] in people with type 2 diabetes have not assessed whether compensatory physical activity, altered diet, or changed sleeping patterns underlie any time-of-day exercise effect. In this 16-week randomised crossover study, we aimed to investigate glycaemic outcomes of moderate intensity exercise at different times of the day in people with type 2 diabetes also being prescribed metformin monotherapy. Secondary outcomes included total physical activity, medication timing, diet, and sleep. Based on previous evidence [2, 5–8], we hypothesised that evening exercise would be more efficacious in lowering glycaemia in the trial.

## 2. Methods

### 2.1. Study Design & Ethical approval

The study was conducted following ethical approval from the UK Health Research Authority’s Integrated Research Application System (IRAS), and via the London Centre Research Ethics Committee (REC reference: 20/PR/0990; IRAS project ID: 292015) and in accordance with the principles of the Declaration of Helsinki as revised in 2008. Informed written consent was obtained from participants during recruitment. The trial was preregistered - Research Registry Unique Identifying Number: researchregistry6311. Nine males and nine females with type 2 diabetes also being prescribed metformin monotherapy completed a remote, randomised, crossover two-arm trial. An overview of the study’s design is shown in Figure 1.

**Figure 1.**
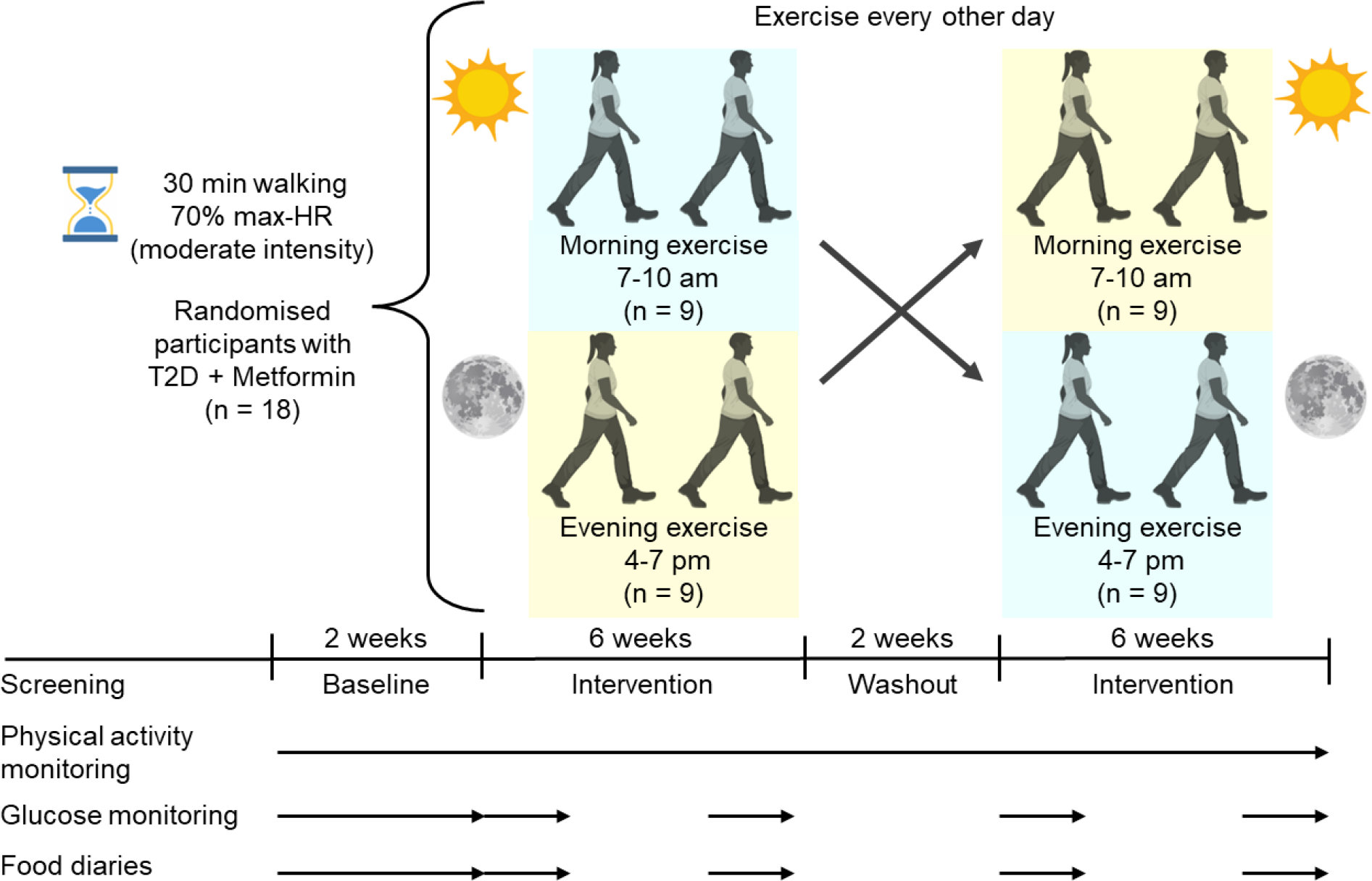
Study Design. HR, Heart rate. Figure created with Biorender.

### 2.2. Participants

#### 2.2.1. Eligibility criteria

To be considered eligible for inclusion, participants had to be diagnosed with type 2 diabetes (insulin independent); have a body Mass Index of 20-36 kg/m^2^; aged 45-75 years, male or female; be prescribed metformin; be able to participate in an exercise intervention; be able to interact with smart-phone apps and be able to provide informed consent. Participants were excluded from the study if they were treated with insulin medication; they were current nicotine users (cigarettes/nicotine gum etc.); they were past nicotine users <6 months before inclusion in the study; have a pre-existing cardiovascular condition; pre-existing blood-borne disease; pre-existing rheumatic illness; have cancer; have a pre-existing psychiatric disorder; another pre-existing systemic disease; extreme chronotype (*e*.*g*., extreme ‘lark’ or ‘owl’); prescribed any glycaemia regulating medications other than metformin; already meeting physical activity guidelines *i*.*e*. > 150 minutes of moderate intensity activity a week or 75 minutes of vigorous intensity activity a week or those who could not adequately read or understand English.

#### 2.2.2. Recruitment

Volunteers were recruited from the Aberdeen and Aberdeenshire areas through the Scottish Primary Care Research Network (SPCRN), over the course of 1 year, between May 2021 and June 2022. The experimental study included recruitment and exercise interventions for two groups, winter, and summer cohort. SPCRN conducted a pre-screening and eligible potential volunteers received a letter of invitation from their National Health Service GP practice. If individuals were interested in participating, they were sent the consent form and questionnaires as part of an information pack. After phone consultation, participants who agreed to continue were instructed to complete, sign and to mail the forms back to the research team. Following a review of the forms to ensure they were properly completed; the necessary equipment was posted to the participants. Subsequently, participants received another phone call from a member of the research team to review the participant information sheet (PIS), answer any questions the participant may have and explain the instructions on how to fit continuous glucose monitors and physical activity monitors.

GPs were notified about their patient’s participation in the study and the researchers were sent participants’ information about BMI, HbA1c, date of type 2 diabetes diagnosis, and time since metformin prescription from the GP records. One participant did not have any available data, five of the participants did not have available records about type 2 diabetes diagnosis date and four participants time since metformin prescription as they were new to the GP.

#### 2.2.3. Sample size calculation

The number of participants (30) was determined in collaboration with an independent statistician at Biomathematics and Statistics Scotland (BioSS) based on our previous study [2]. *i*.*e*. an estimated difference in mean glucose concentration of 0.69 mmol/L, an estimated standard deviation (SD) of 1.32 mmol/L (SD of the individual differences between two exercise treatments), a paired analysis, a significance level of 0.05 and a power of 80%.

#### 2.2.4. Randomization to morning or evening exercise

Randomisation was carried out by an independent statistician at BioSS. Microsoft Excel was used to randomly allocate participants to the two possible sequences (*i*.*e*., morning first or evening first) in blocks by order of joining the trial and sex. The randomisation was generated once we received a positive response from the participant.

#### 2.2.5. Adherence monitoring

Compliance with the exercise regimen were assessed using real-time data obtained from the Garmin Connect (Garmin Ltd, Olathe, KS, US) and LibreView (Abbott Diabetes Care Inc, Alameda, CA, US) sites. Participants missing pre-defined adherence targets (*i*.*e*., either missing >4 exercise windows in less than 2 weeks, with a heart rate discrepancy of >15% during exercise or >4 days with no glucose reading) were contacted by phone call or e-mail and encouraged to improve adherence. If adherence was not improved, they were not considered for the final data analysis. When the trial coincided with the Christmas holidays, the wash-out period was extended for a period of 4 weeks so that the intervention arms did not run during the Christmas holidays.

### 2.3. Exercise Intervention

Participants were requested to perform 30 minutes of walking every other day during a 6-week period, maintaining, as close as possible, 70% of their estimated maximum heart rate (estimated with the formula 220 minus age [14]) during exercise. Participants were instructed to perform exercise within a 3-hour window between either 7:00-10:00 AM or 4:00-7:00 PM. The overall 16-week trial period included 2-week baseline recording, 12-week intervention arms (6 weeks morning and then 6 weeks evening exercise, or vice versa) with a 2-week washout between trial arms. Physical activity data were collected for the entirety of the trial, pre-trial and wash-out period.

### 2.4. Outcomes

#### 2.4.1. Physical activity

Physical activity data (assessed as step count per day and heart rate) as well as sleep quality were collected via a wrist-based physical activity monitor Garmin Vivosmart 4 (Garmin Ltd, Olathe, KS, US), which participants were asked to wear for the entire duration of the trial. Participants were instructed to download the Garmin Connect application and log in with a study password. The outcomes of the physical activity monitors were step count per day, heart rate and sleep quality. Participant’s chronotype was determined using the self-report Munich Chronotype Questionnaire.

#### 2.4.2. Glucose monitoring

Participants were provided with continuous glucose monitors The FreeStyle Libre 2 sensor (CGMs, Abbott Diabetes Care Inc, Alameda, CA, US), that measures the glucose concentration in interstitial fluid continuously, storing a reading every 15 minutes. Sensors had a working life of 2-weeks. Glycaemia data was collected by the LibreView (Abbott Diabetes Care Inc, Alameda, CA, US) cloud system, participants were instructed to download the FreeStyle LibreLink application on their own smart phones to share and link their data to a LibreView account in real time. Glucose information was collected for 2 weeks before start of the intervention, during the first 2 weeks, and for the last two weeks of each exercise intervention.

#### 2.4.3. Dietary Intake and metformin dose

Participants were asked to maintain their normal diet consistently throughout the trial and record their diet throughout the trial using a 4-day food diary in which the participant recorded the food eaten, the time of day each meal was consumed, the cooking methods used, and the portion sizes. A 4-day food diary was completed during the baseline period, and during the first 2 weeks, and the last 2 weeks of each exercise intervention. Dietary intake was determined using Windiet software (Univation Ltd, The Robert Gordon University, Aberdeen, Scotland, UK). Participants were also asked to record the time and dose of metformin on their food diaries. As dietary intake and metformin time and dose were self-reported, the data analysis of the food diaries and metformin time-doses corresponded to 14 participants and the metformin analysis is from 13 participants that fulfilled this requirement.

### 2.5. Data analysis

Blood glucose levels were measured using CGMs, from which 24-hours hourly mean were calculated using the R Studio statistical software package v2022.12 (Rstudio.com; R version 4.2.2, R Foundation for Statistical Computing, Boston, MA, US). Data was then analysed using Prism 9.5.1 software (GraphPad, San Diego, CA, US) by One-way ANOVA or two-way ANOVA followed up by Holm-Šídák’s multiple comparisons test (significance level set at *p*<0.05). Baseline characteristics between males and females were compared by means using t-test (significance level set at *p*<0.05). Results are expressed as Mean±SEM and estimates with 95% CIs.

## 3. Results

### 3.1. Participants recruitment and characteristics

Of the forty positive responses received, nine male and nine females with type 2 diabetes being prescribed metformin completed the 16-week crossover intervention; see Figure 2 for consolidated Standards of Reporting Trials (CONSORT) flow chart. Following randomization, eight participants withdrew for personal reasons, four of them from the winter cohort and four from summer group. After completion of the trial, five participants did not meet the exercise intervention requirements, finishing with eleven participants from winter; and seven from the summer cohort. In terms of crossover sequence, the study finished with eight participants who did morning exercise first and ten participants who did evening exercise first. Sleep data corresponds to fifteen participants who fulfilled this requirement, glucose data was missing from one participant during the last 2 weeks of the morning period and from two participants during the last 2 weeks of evening exercise. Baseline characteristics of study participants that completed the trial are shown in Table 1 (Characteristics according to different stratification are shown in supplementary Tables 1,2,3,4,5).

**Table 1.** Baseline characteristics of study participants.

| Characteristic (n = 18) | Mean ± SEM |
| --- | --- |
| Sex | Male n=9; Female n=9 |
| Age (years) | 61±2 |
| BMI (Kg/m<sup>2</sup>) | 30.6±0.7 |
| HbA1c (mmol/mol) | 64.4±15.0 |
| HbA1c (%) | 8.0±0.3 |
| Time since T2D diagnosed (years) | 8.2±1.7 |
| Dose of Metformin per day (mg) | 1313.0±134.6 |
| Time of Metformin (years) | 5.2±1.0 |
| Steps per day at baseline | 6798±595 |

*T2D, Type 2 Diabetes*

**Figure 2.**
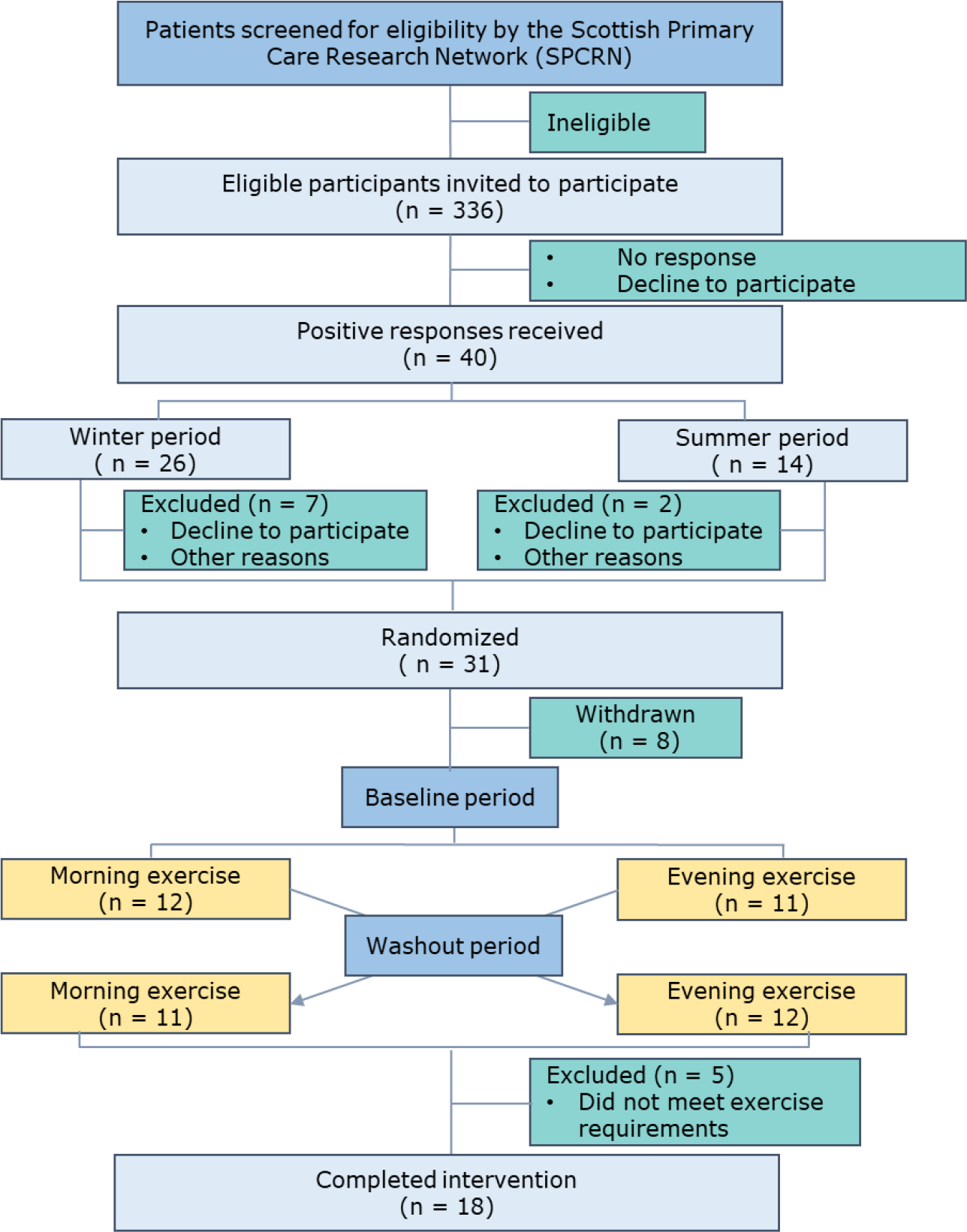
Consolidated Standards of Reporting Trials (CONSORT) flow chart.

### 3.2. Adherence to exercise intervention

The total number of exercise sessions participants were asked to complete during each exercise period was 21. The exercise completion rate was consistent (*p*=0.47) between morning (19.1±1.3 sessions) and evening exercise (18.1±1.4 sessions) (Supplementary Figure 1). Total physical activity was similar between the arms of the trial (Figure 3A, B). The number of mean daily steps recorded during baseline was 6798.0±595.6. During exercise days participants completed a mean daily total of 10814.0±530.7 steps and 10373.0±514.6 steps during morning and evening, respectively (*p*=0.15). During rest days participants completed a mean daily total of 6843.0±561.8 steps and 6344.0±514.3 steps during morning and evening, respectively (*p*=0.24). Total hours of sleep registered from Garmin devices (8.3±0.3) was significantly higher (*p*=0.03) the hours of sleep recorded from self-reported MCTQ (7.67±0.2) (Supplementary Figure 2). The total hours of sleep indicated a trend (one-way ANOVA: *p*=0.06) showing longer sleep duration during evening exercise (8.3±0.2 hours) compared with morning exercise (7.9±0.2 hours), however there was no difference compared with baseline (*p*=0.31, *p*=0.40, morning exercise, evening exercise, respectively) (Figure 3C). Sleep architecture such as deep sleep, light sleep, REM sleep and time awake showed no significant difference between baseline, morning and evening exercise (*p*=0.35, *p*=0.57, *p*=0.06, *p*=0.20, respectively) (Supplementary Figure 3). Participants were instructed to exercise at an estimated 70% of max-HR, which was 111.4±1.3 bpm. Exercise intensity during morning and evening exercise was similar (Figure 3D), as measured by HR (117.2±1.9 bpm and 117.3±2.7 bpm, respectively, p=0.93). Overall, our results show no differences in physical activity or compensatory activity between morning and evening exercise.

**Figure 3.**
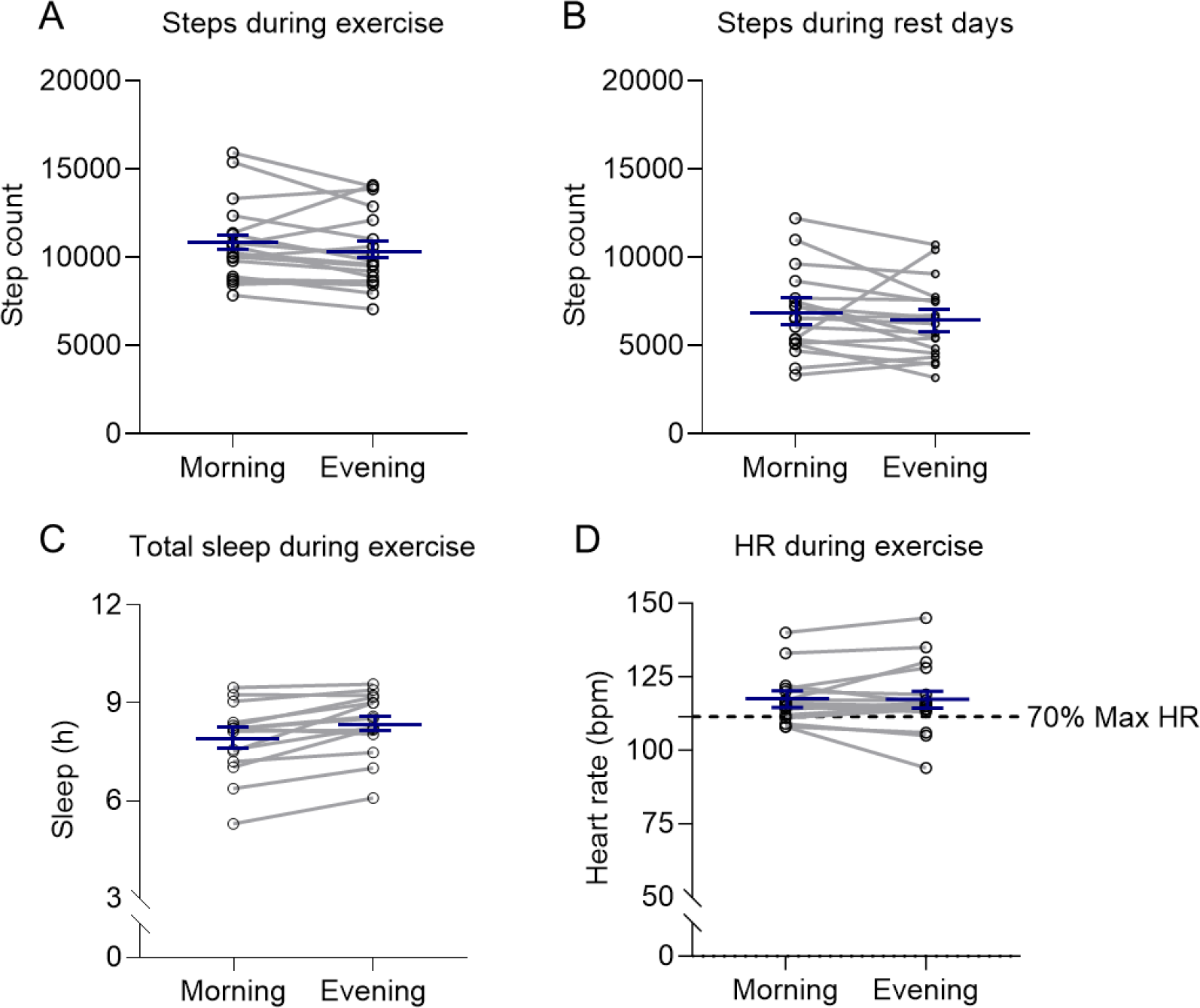
Step count, max-HR and total sleep hours during the morning and evening exercise periods. The values are shown for number of steps on (A) Days when the participants performed exercise (n=18) and (B) Rest days (n=18); (C) Total sleep hours during exercise (n=15) and (D) Max-HR (n=18) throughout trial; blue lines are mean±SEM, dotted line (Figure D) represents 70% Max-HR estimated; HR, Heart rate. Step count, total hours of sleep and sleep insights were analysed using one-way ANOVA, total hours of sleep from Garmin devices vs MCTQ and Max-HR were analysed using paired t-test.

### 3.3. Changes in glucose during exercise intervention

There was a significant decrease (*p*=0.02) in acute (first 24 hours of exercise) AUC glycemia during morning exercise (180.6±16.1 mmol/L) compared with baseline (210.3±18.0 mmol/L), whereas there were no significant differences in acute glycemia levels during evening exercise compared with baseline (*p*=0.12). No change in acute glucose levels between morning and evening exercise were observed (*p*=0.14) (Figure 4A). An hour-by-hour time-course of glucose readings did not indicate a difference in glycemia during the trial (*p*>0.05) (Figure 4B).

**Figure 4.**
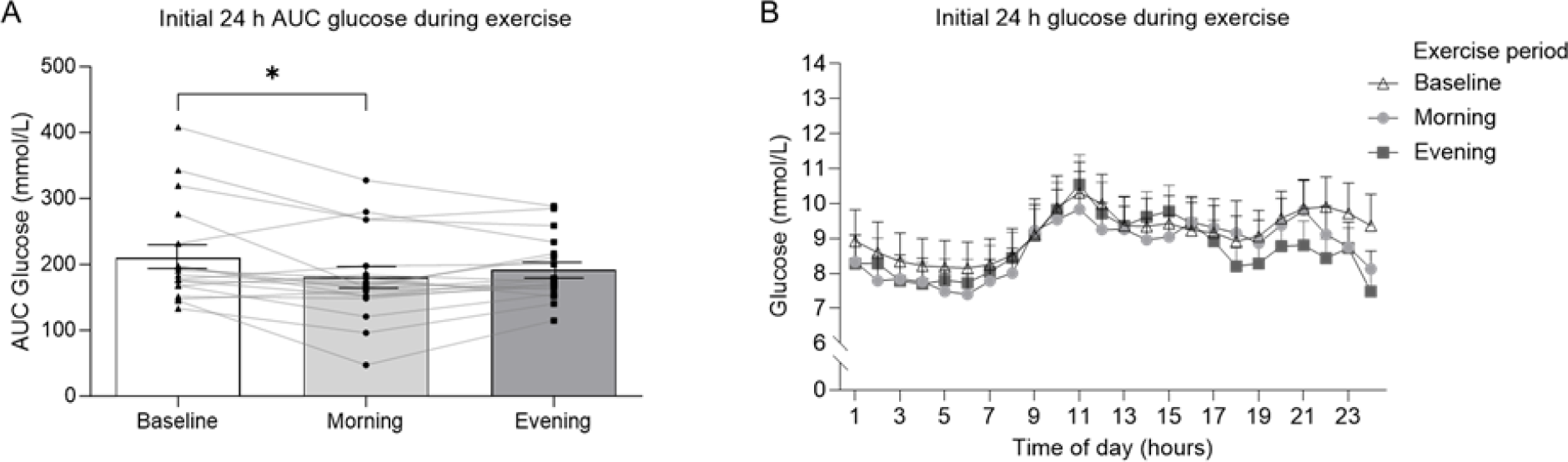
Acute 24-hour glucose levels. (A) Area under the curve (AUC) values for glucose levels during the first 24 hours of exercise. (n=18) (B) Time-course plot hourly glucose levels for the first 24 hours of exercise during baseline, morning, and evening intervention (n=18). Wk, week. *p< 0.05. Values are mean±SEM, lines represent individual values. Data were analysed using one-way ANOVA followed up by Holm-Šídák’s multiple comparisons test.

24-hour AUC glucose during baseline (210.3±18.0 mmol/L), morning (197.3±13.9 mmol/L) and evening (206.7±14.9 mmol/L) during weeks 1-2 did not differ significantly (*p*=0.24) (Figure 5A). Further, no difference in glucose concentration was observed at any time point during the 24-hour time-course between baseline, morning, and evening exercise, during weeks 1-2 (Figure 5B). Similar findings were observed at weeks 5-6 (Figure 5D).

**Figure 5.**
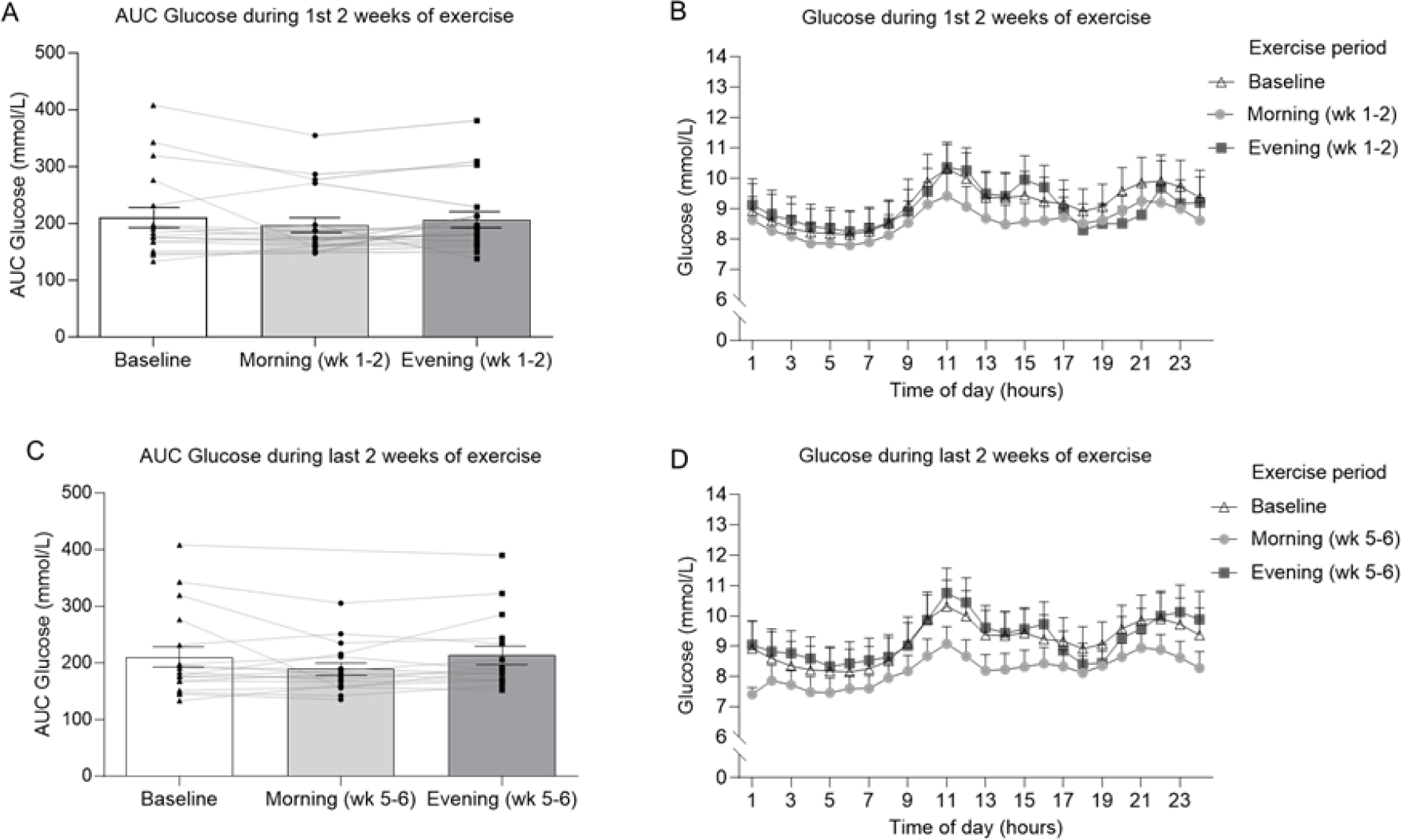
24-hour glucose levels in response to exercise intervention. (A) Area under the curve (AUC) 24-hour glucose for week 1-2 of exercise intervention (n=18) (B) Time-course plot 24-hour glucose in weeks 1-2 (n=18) (C) Area under the curve (AUC) 24-hour glucose for week 5-6 of exercise intervention (n=17) (D) Time-course plot 24-hour glucose in weeks 5-6 (n=17). Wk, week. Values are mean±SEM, lines represent individual values. Data were analysed using one-way ANOVA followed up by Holm-Šídák’s multiple comparisons test.

No significant interaction effect was observed for sex x time of the exercise (Supplementary Figure 4). There was a trend (*p*=0.07) showing lower AUC glucose levels in summer (222.4±6.1) compared to the winter (169.8±0.9). This consistent pattern persisted throughout the entire intervention period, however, this difference in AUC glucose levels between winter and summer did not reach statistical significance (*p*=0.07) (Supplementary Figure 5).

### 3.4. Caloric intake during exercise intervention

There were no changes in dietary energy intake during breakfast (*p*=0.30), baseline (390.5±38.2 Kcal), morning weeks 1-2 (355.0±22.8 Kcal), evening weeks 1-2 (353.1±49.6 Kcal), morning weeks 5-6 (311.6±27.21 Kcal), evening weeks 5-6 (354.2±40.71 Kcal) throughout the trial (Figure 6A). Energy intake during lunch was significantly different (*p*=0.03) between morning weeks 1-2 (534.3±61.0 Kcal) and evening weeks 5-6 (402.7±45.5 Kcal) (Figure 6B). However, no significant differences were observed (*p*=0.64) when comparing evening weeks 5-6 with the energy intake at baseline (470.4±41.3 Kcal). There was no difference in caloric ingestion of dinner (*p*=0.14) (Figure 6C) between arms of the trial. No changes in caloric intake were detected between comparable periods of the trial.

**Figure 6.**
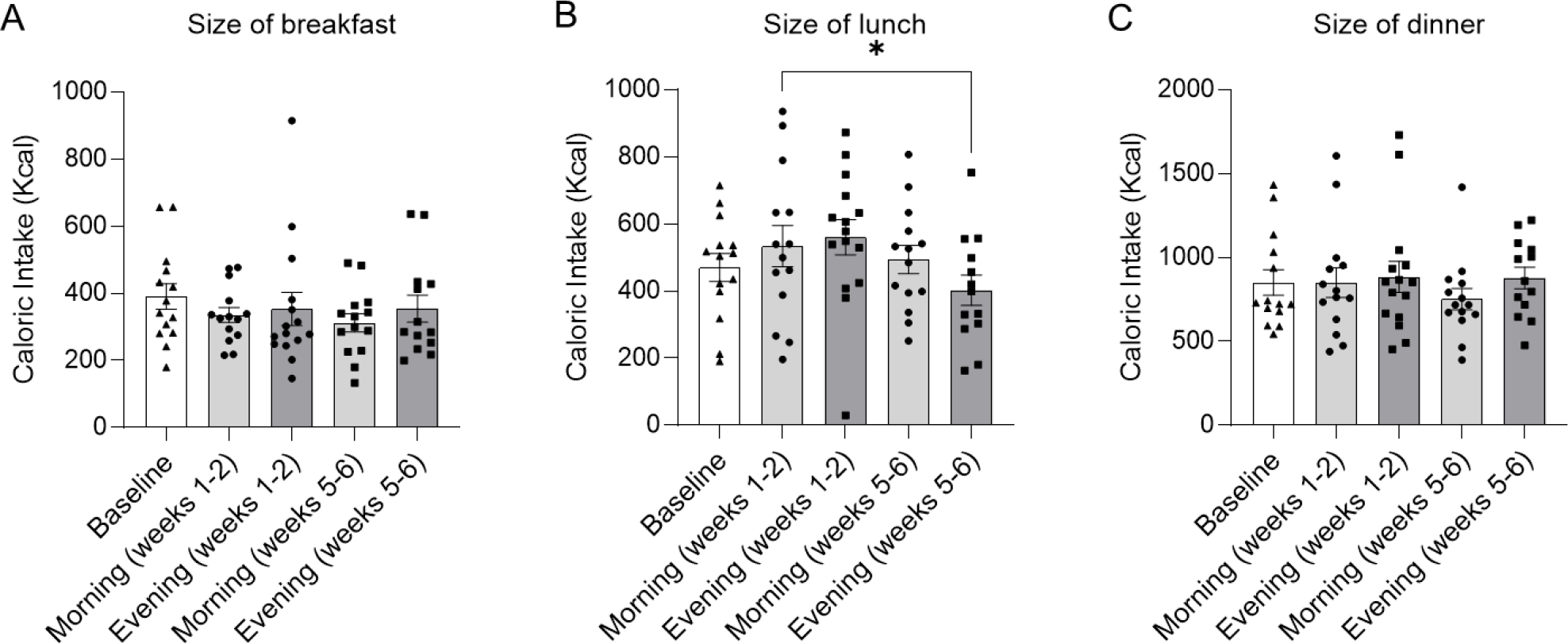
Mean caloric intake during exercise intervention. Data collected from self-reported 4-days food diaries for (A) Size of breakfast (n=14), (B) Size of lunch (n=14) and (C) Size of dinner (n=14). Values are mean±SEM. Data were analysed using one-way ANOVA followed up by Holm-Šídák’s multiple comparisons test.

There were no differences (p=0.33) in total daily caloric intake consumed during baseline (570.6±142.1 Kcal), morning weeks 1-2 (573.3±149.9 Kcal), evening weeks 1-2 (533.1±116.8 Kcal), morning weeks 5-6 (585.2±165.6 Kcal) and evening weeks 5-6 (545.2±167.3 Kcal) (Figure 7). Throughout the trial, caloric intake was significantly higher (p=<0.001) during dinner (843.1±23.8 Kcal), compared with breakfast (349.0±13.0 Kcal) and lunch (492.4±27.3 Kcal).

**Figure 7.**
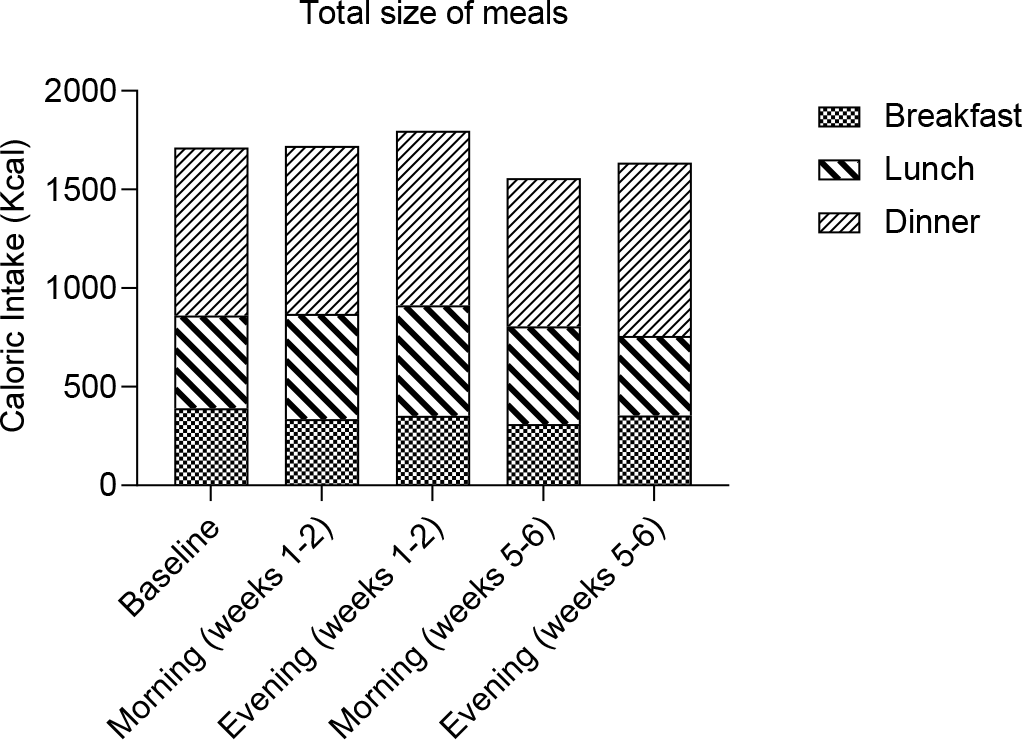
Total daily caloric intake. Data collected from self-reported 4-days food diaries from baseline (n = 14), morning weeks 1-2 (n = 14), evening weeks 1-2 (n=15), morning weeks 5-6 (n=14), evening weeks 5-6 (n=13). Values are mean±SEM. Data were analysed using one-way ANOVA followed up by Holm-Šídák’s multiple comparisons test.

### 3.5. Meals and metformin timing

Acute 24 AUC glucose was lower (*p*=0.01) in participants taking metformin before breakfast (152.5±10.6 mmol/L) compared with participants taking metformin after breakfast (227.2±27.5 mmol/L) only when the participants performed morning exercise (Figure 8B). No differences were observed for metformin before or after breakfast for the baseline and evening exercise arms of the trial (*p*=0.17 and *p*=0.67, respectively), nor for metformin before or after dinner during baseline, or morning or afternoon exercise (*p*=0.44 and *p*=0.25, respectively) (Supplementary Figure 6).

**Figure 8.**
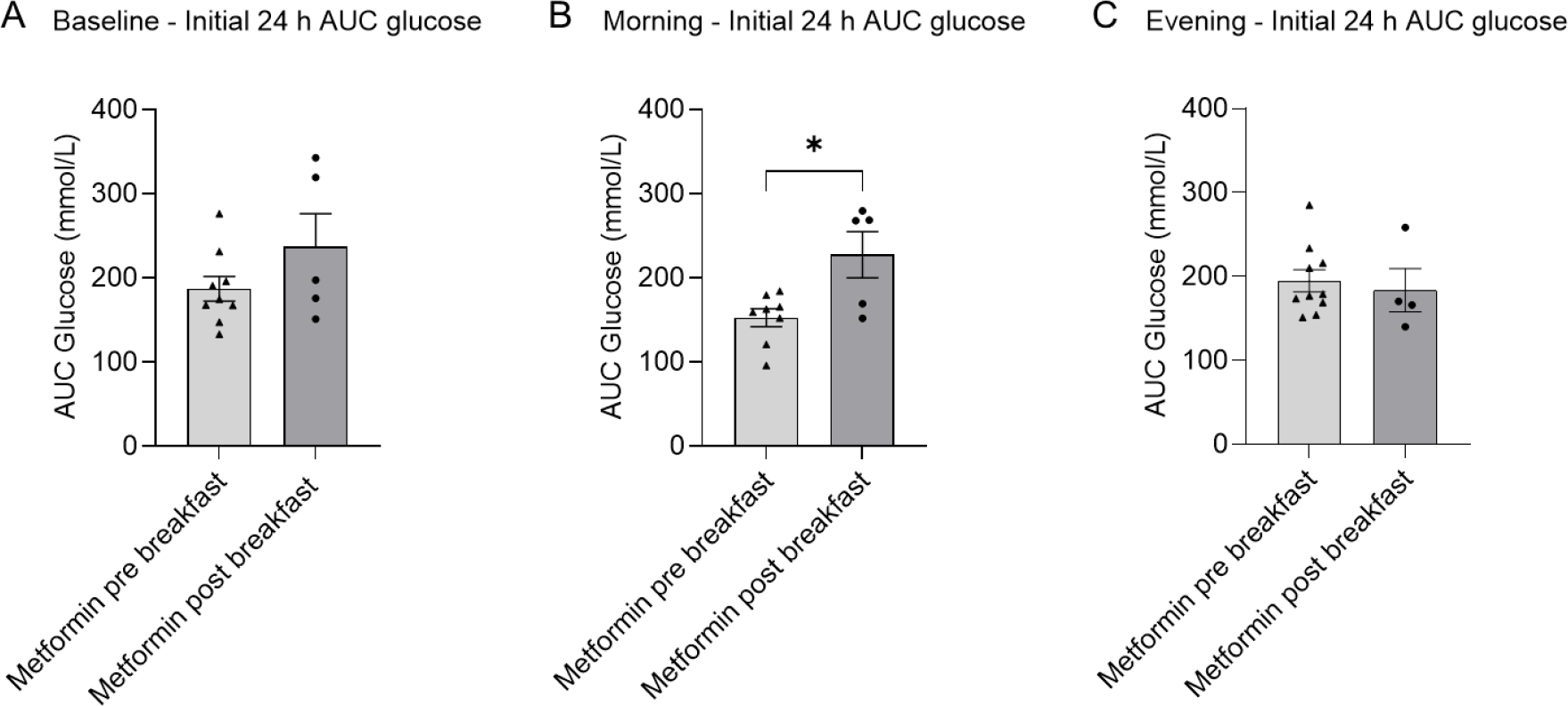
Meal and metformin timing. Area under the curve (AUC) values for glucose levels during the first 24 hours of exercise for (A) Baseline, metformin pre-breakfast (n=9), metformin post-breakfast (n=5); (B) Morning exercise, metformin pre-breakfast (n=8), metformin post-breakfast (n=5); (C) Evening exercise, metformin pre-breakfast (n=10), metformin post-breakfast (n=4). *p< 0.05. Values are mean±SEM. Data were analysed using unpaired t-test.

During morning exercise (weeks 1-2), participants taking metformin before breakfast exhibited a trend (p=0.07) to have lower AUC glucose levels (165.4±3.3 mmol/L) compared with participants taking metformin after breakfast (223.3± 25.1 mmol/L) (Figure 9). During weeks 5-6 of the exercise protocol, glucose was significantly lower (p=0.04) for participants taking metformin before breakfast (168.8±5.6), rather than after breakfast (224.5±21.2) only during morning exercise. No differences in AUC glucose for metformin before/after breakfast were observed for the evening arm of the trial throughout the protocol (evening arms, weeks 1-2, p=0.80, weeks 5-6 p=0.72).

**Figure 9.**
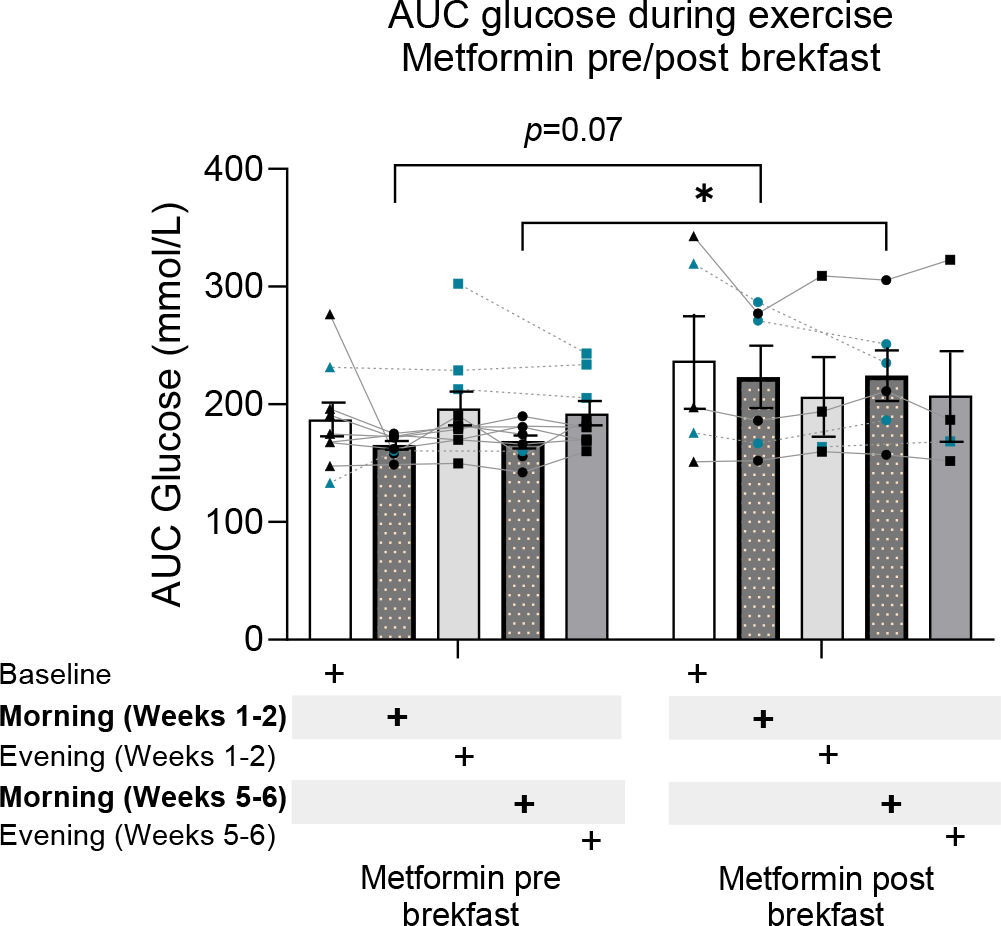
Meal and metformin timing throughout the trial. Area under the curve (AUC) values for glucose levels for Baseline period, metformin pre-breakfast (n=9), metformin post-breakfast (n=5); Morning exercise (weeks 1-2), metformin pre-breakfast (n=8), metformin post-breakfast (n=6); Evening exercise (weeks 1-2), metformin pre-breakfast (n=10), metformin post-breakfast (n=4); Morning exercise (weeks 5-6), metformin pre-breakfast (n=8), metformin post-breakfast (n=6); Evening exercise (weeks 5-6), metformin pre-breakfast (n=9), metformin post-breakfast (n=4). *p< 0.05. Values are mean±SEM. Blue dots are participants that changed metformin intake timing between different periods of the trial, lines represent individual crossover values. Data were analysed using two-way mixed-model ANOVA followed up by Holm-Šídák’s multiple comparisons test.

## 4. Discussion

These data demonstrate that morning moderate intensity exercise acutely reduces glycaemia in people with type 2 Diabetes also being prescribed metformin. While evening exercise had no effect upon glycaemia. The acute reduction of glycaemia in the morning trial was apparently driven by people who consumed metformin before breakfast, rather than after breakfast. Indeed, morning exercise combined with pre-breakfast metformin persistently reduced AUC glucose compared to morning exercise combined with post-breakfast metformin up to the final week (week 6) of the intervention. In this crossover trial, there was no significant difference in exercise intensity, caloric intake or total physical activity between the arms of the trial (morning vs evening exercise). These findings support the assertion that an interaction with intrinsic circadian biology may be linked to the observed effects.

Interestingly, we observed that the acute morning exercise decrease in glycaemia was apparently driven by people who consumed metformin before breakfast rather than after breakfast. Many people taking metformin struggle to maintain regular physical activity [15], which may be a result of a lower tolerance of exercise when metformin is concomitant [16]. Furthermore, metformin may interfere with the glucose-lowering effect of acute exercise [11, 12]. Therefore, identifying strategies to optimise concomitant prescription of these therapies is essential. Our data suggests that it may be possible to optimise timing of these concomitant therapies to augment their therapeutic effect. It should be noted that these specific data in the current study are not crossover, however, the magnitude of difference (33% decreased AUC glucose in pre-breakfast group, Figure 8) suggests a real effect. Additionally, an interaction effect on glycaemia between morning exercise and pre-breakfast metformin intake appeared to persist throughout the trial since the group taking metformin before breakfast had lower AUC glucose than the post breakfast group into week 6 of the morning exercise intervention. These findings have parallels to previous data that suggest consuming metformin before a meal improved efficacy upon glycaemic regulation [17]. This effect on glycaemia may be partly related to metformin’s pharmacokinetic interaction with meal intake and intrinsic circadian rhythms. For example, compared with the fasting state, bioavailability of metformin is 24% lower, and the peak concentration delayed about 37 min when an 850 mg tablet is administered with food [18]. Additionally, metformin’s pharmacology significantly depends on time-of-day in humans, which may be related to glomerular filtration rate, renal plasma flow (RPF) and renal organic cation transporter (OCT) 2 activity [19]. Although the half-life of metformin in the blood is relatively long (∼18 hours depending on dose/method [20]), less is known regarding accumulation in skeletal muscle, a key organ in the response to exercise [1]. It is known that concomitant exercise alters the pharmacokinetics of acute metformin administration [16, 20], with differing skeletal muscle concentrations of metformin before, during and after exercise [16]. It is plausible then, that exercise at different times of the day in addition to timing of metformin intake may interact with the pharmacokinetics and glycaemic modulating effects of metformin. This is supported by our findings since we observed a glucose lowering effect of pre-breakfast metformin only during morning exercise periods of the trial. Thus, further research should aim to fully elucidate the interaction between meal-timing, exercise and medication intake.

As described above, skeletal muscle plays a key role in the metabolic response to exercise, and this organ has an intrinsically disrupted rhythm in people with type 2 Diabetes [9]. However, it is plausible that metformin treatment may interact with skeletal muscle circadian metabolism to alter time-of-day exercise effects observed in other studies in people with impaired metabolism. Supportive of this is the lack of effect upon glycaemia in response to exercise at different times of the day in people who are metabolically healthy [10] and who presumably do not have disrupted metabolic circadian rhythms. However, the aforementioned study [10] only monitored response to a single bout of exercise, while the current study was a 16-week trial in which participants did two 6-week training periods. Moderate intensity exercise did not increase glycaemia in people with type 2 Diabetes being prescribed metformin. These data indicate that in terms of glycaemic response, morning moderate intensity exercise is not deleterious in people with type 2 diabetes being prescribed metformin and does not exacerbate the dawn phenomenon in the same manner as HIIT [2].

Indeed, exercise modality may interact with circadian rhythm to alter exercise outcomes. High-intensity exercise capacity is consistently higher in the afternoon/evening compared to morning, while diurnal differences in exercise capacity are less clear with moderate intensity exercise [21]. This phenomenon may underlie the findings of Teo et al. (2020), who found no time-of-day differences in glycaemic outcomes in people with type 2 diabetes in response to combined walking exercise and resistance training [13]. This exercise modality [13] contrasts with the HIIT protocol used in previous studies with conflicting findings [2]. However, it should be noted that Teo et al. (2020) did not measure glycaemic outcomes in a circadian manner [13]. Whereas, in the current study we have measured 24-hour glucose over a total period of 10-weeks. Additionally, participants completed moderate intensity exercise outdoors in

Scotland, which may influence diurnal outcomes compared to exercise protocols completed at a higher intensity or in a different environment. In a laboratory setting, fifty minutes of walking at 3 different times of day and at different timing in relation to meals did not lower 24-hour glucose concentrations in people with type 2 diabetes [22]. Together with the results from the current study, these data suggest that moderate intensity exercise may not give rise to time-of-day dependent glycaemic perturbations of the same magnitude as high intensity exercise.

The current study had some limitations. Our prior statistical power calculations suggested a sample size of 30 in order to detect differences in hour-by-hour glucose readings. Although we met our recruitment target, we had a higher-than-expected withdrawal rate, with 18 participants eligible for final analysis. We conducted this study mainly during the C-19 global pandemic, which directly and indirectly impacted on participants’ ability to complete the 16-week trial. Nevertheless, the strength of our study is the cross-over design, giving greater statistical power than a cross-sectional design. We aimed to recruit people with type 2 diabetes who were being prescribed metformin monotherapy, *i*.*e*. no other glycaemic regulating pharmaceutical treatments. This criterium allowed us to investigate a time-of-day exercise interaction with metformin treatment specifically, rather than being confounded by other pharmaceutical treatments. As previously noted, metformin timing intake was not a primary outcome of this study, and therefore our comparisons between different metformin intake timings are not crossover. Thus, it is possible that existing differences in glycaemic regulation between groups are responsible for some of the observed differences (*e*.*g*. Supplementary table 4). However, it is likely that metformin intake timing is highly habitual, and it is plausible that participants who habitually take metformin before breakfast may experience better long-term control of glycaemia as a direct result. A further strength of the study was the excellent adherence of participants to the exercise protocol, with >85-90% exercise session completion rate over 6-weeks and no difference in self-monitored exercise intensity between arms of the trial. The advantage of conducting self-monitored exercise trials is that this is likely to better recapitulate many real-world settings in which clinicians’ recommendations to exercise are mostly self-monitored rather than monitored by practitioners. Participants were instructed to maintain their normal diet during the trial, and total caloric intake was not different between arms of the trial, indicating adherence to this instruction. However, it should be noted that dietary intake was self-reported.

In summary, our study demonstrates an acute decrease in glycaemia in response to morning exercise in people with type 2 diabetes also being prescribed metformin. The reduction of glycaemia was driven by participants who consumed metformin before breakfast, rather than after breakfast, indicating an interaction between meal-timing, metformin intake and exercise. In this trial we monitored diet, exercise intensity, and physical activity throughout the trial, none of which appeared to contribute to the time-of-day dependent effect of exercise on glycaemia. Many people being prescribed metformin struggle to maintain regular physical activity [15], and metformin may interfere with the glucose-lowering effect of acute exercise [11, 12]. Thus, finding strategies to augment the therapeutic effect of these treatments when concomitant is essential in the management of type 2 diabetes. Our research indicates that it may be possible to optimise the timing recommendations for concomitant exercise and metformin treatment. Specifically, our findings suggest that morning moderate intensity exercise combined with pre-breakfast metformin intake may benefit the management of glycaemia in people with type 2 diabetes.

## Supporting information

CONSORT_checklist

## Abbreviations

BIOSS: Biomathematics and Statistics Scotland
CGMs: Continuous glucose monitors
CONSORT: Consolidated Standards of Reporting Trials
GP: General Practitioner
HIIT: High intensity interval training
IRAS: Integrated Research Application System
MCTQ: Munich Chronotype Questionnaires
OCT: Renal organic cation transporter
REC: Research Ethics Committee
REM: Rapid eye movement
RPF: Renal plasma flow
SEM: Standard error of mean
SPCRN: Scottish Primary Care Research Network
Max-HR: Maximum heart rate

## 5. Acknowledgements

We would like to thank Amanda Cardy from the NRS Primary Care Network for her help in recruiting participants. We would also like to thank Dr Graham Horgan of Biomathematics & Statistics Scotland (BIOSS) for his support with statistical analysis.

## 7. Data availability

Raw data underlying all figures in this manuscript have been deposited publicly: 10.6084/m9.figshare.24081417

