## Supplementary Data for "Morning exercise and pre-breakfast metformin interact to reduce glycaemia in people with Type 2 Diabetes: a randomized crossover trial"

**SUPPLEMENTARY MATERIAL**

**Supplementary table 1**. Baseline characteristics by gender. HbA1c, blood glycosylated haemoglobin. Values are mean ± SEM (n). *P* values are from unpaired t-test.

| Characteristic | Males *(n)* | Females *(n)* | *p* value |
|---|---|---|---|
| Age (years) | 58.3±2.7 (9) | 63.6±2.8 (8) | 0.20 |
| BMI (Kg/m$^2$) | 31.4±1.0 (9) | 29.8±1.0 (8) | 0.28 |
| HbA1c (mmol/mol) | 65.6±5.6 (9) | 63.1±4.9 (8) | 0.74 |
| HbA1c (%) | 8.1±0.5 (9) | 7.9±0.4 (8) | 0.71 |
| Time since T2D diagnosed (years) | 3.6±0.9 (5) | 6.6 ±1.6 (7) | 0.26 |
| Dose of Metformin per day (mg) | 1500.0±288.7(3) | 1250±155.9(9) | 0.45 |
| Time of Metformin (years) | **4.0±1.0 (5)** | **11.4±2.4 (8)** | **0.04** |
| Number of steps | 7114±984.4 (9) | 6481±713.2 (9) | 0.60 |

**Supplementary table 2**. Baseline characteristics by season. HbA1c, blood glycosylated haemoglobin. Values are mean ± SEM (n). P values are from unpaired t-test.

| Characteristic | Winter *(n)* | Summer *(n)* | *p* value |
|---|---|---|---|
| Age (years) | 60.5±2.3 (10) | 61.29±3.87 (7) | 0.86 |
| BMI (Kg/m$^2$) | 30.4±0.9 (10) | 31.0±1.3 (7) | 0.69 |
| HbA1c (mmol/mol) | **70.7±4.9 (10)** | **55.6±3.5 (7)** | **0.04** |
| HbA1c (%) | **8.7±0.4 (10)** | **7.3±0.3 (7)** | **0.03** |
| Time since T2D diagnosed (years) | 4.6±2.2 (5) | 5.8±1.0 (7) | 0.59 |
| Dose of Metformin per day (mg) | **1042.0±100.3(6)** | **1583.0±200.7(6)** | **0.04** |
| Time of Metformin (years) | 10.3±3.5 (6) | 7.0±1.6 (7) | 0.38 |
| Number of steps | 6184±655.9 (11) | 7761±1094 (7) | 0.20 |

**Supplementary table 3**. Baseline characteristics by metformin timing during baseline period. HbA1c, blood glycosylated haemoglobin. Values are mean ± SEM (n). P values are from unpaired t-test.

| **Baseline period** | | | |
|---|---|---|---|
| **Characteristic** | **Metformin before breakfast *(n)*** | **Metformin after breakfast *(n)*** | ***p* value** |
| Age (years) | 59.1±2.6 (9) | 64.3±4.0 (4) | 0.30 |
| BMI (Kg/m$^2$) | 30.6±1.3 (9) | 30.8±1.1 (5) | 0.91 |
| HbA1c (mmol/mol) | 56.6±4.6 (9) | 68.8±6.6 (5) | 0.26 |
| HbA1c (%) | 7.6±0.4 (9) | 8.4±0.6 (5) | 0.27 |
| Time since T2D diagnosed (years) | 8.4±2.5 (9) | 10.7±0.9 (3) | 0.64 |
| Dose of Metformin per day (mg) | 1918.0±511.3(8) | 1333.0±333.3(3) | 0.52 |
| Time of Metformin (years) | 6.4±1.2 (8) | 3.7±2.7 (3) | 0.29 |
| Number of steps | 7396±1025 (9) | 6133±801.5 (5) | 0.42 |

**Supplementary table 4**. Baseline characteristics by metformin timing in the morning exercise period. HbA1c, blood glycosylated haemoglobin. Values are mean ± SEM (n). P values are from unpaired t-test.

| **Morning exercise** | | | |
|---|---|---|---|
| **Characteristic** | **Metformin before breakfast *(n)*** | **Metformin after breakfast *(n)*** | ***p* value** |
| Age (years) | 59.1±2.9 (8) | 63.4±3.5 (5) | 0.86 |
| BMI (Kg/m$^2$) | 30.8±1.4 (8) | 30.8±1.1 (5) | 0.99 |
| HbA1c (mmol/mol) | **56.9±4.3 (8)** | **75.2±4.7 (5)** | **0.01** |
| HbA1c (%) | **7.3±0.4 (6)** | **9.0±0.4 (5)** | **0.02** |
| Time since T2D diagnosed (years) | 5.6±1.0 (7) | 4.7±3.7 (3) | 0.74 |
| Dose of Metformin per day (mg) | 1357.0±179.8(7) | 1167±166.7(3) | 0.54 |
| Time of Metformin (years) | **10.3±3.5 (8)** | **7.0±1.6 (3)** | **0.02** |
| Number of steps | 7056±1096 (8) | 6752±1141 (5) | 0.86 |

**Supplementary table 5**. Baseline characteristics by metformin timing in the evening exercise period. HbA1c, blood glycosylated haemoglobin. Values are mean ± SEM (n). P values are from unpaired t-test.

| **Evening exercise** | | | |
|---|---|---|---|
| **Characteristic** | **Metformin before breakfast *(n)*** | **Metformin after breakfast *(n)*** | ***p* value** |
| Age (years) | 60.9±1.8 (8) | 61.0±8.7 (3) | 0.98 |
| BMI (Kg/m$^2$) | 30.5±1.3 (8) | 32.6±1.5 (3) | 0.41 |
| HbA1c (mmol/mol) | 66.7±5.8 (8) | 60.3±7.6 (3) | 0.55 |
| HbA1c (%) | 8.2±0.5 (8) | 7.7±0.7 (3) | 0.55 |
| Time since T2D diagnosed (years) | 11.0±3.4 (6) | 9.0±2.5 (3) | 0.71 |
| Dose of Metformin per day (mg) | 1333.0±166.7(6) | 1333.0±333.3(3) | 0.99 |
| Time of Metformin (years) | **8.2±1.0 (8)** | **2.0±1.0 (3)** | **0.008** |
| Number of steps | 6264±758.3 (8) | 5603±1119 (3) | 0.65 |

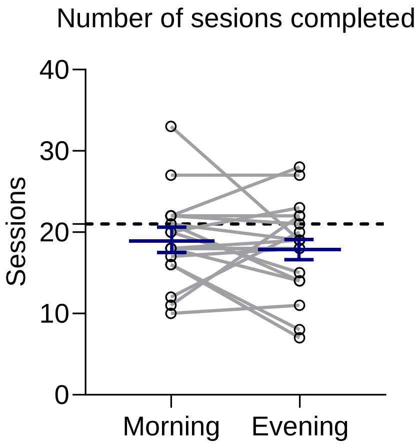


**Supplementary Figure 1.** Number of exercise sessions completed during trial in the morning and evening exercise periods (n=18), blue lines are mean±SEM, the dotted line represents the number of sessions participants were asked to complete. Data were analysed using paired t-test.

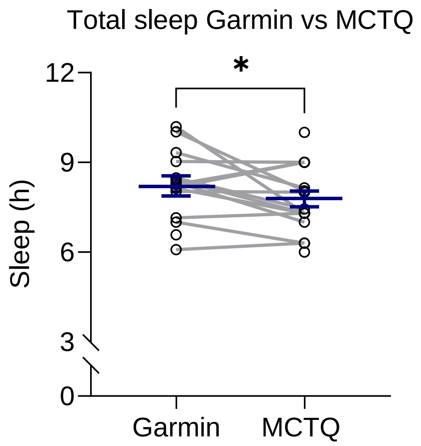


**Supplementary Figure 2.** Total hours of sleep from Garmin devices at baseline (Garmin) and Munich Chronotype Questionnaires (MCTQ) (n=14), blue lines are mean±SEM, data were analysed using paired t-test. *=p<0.05.

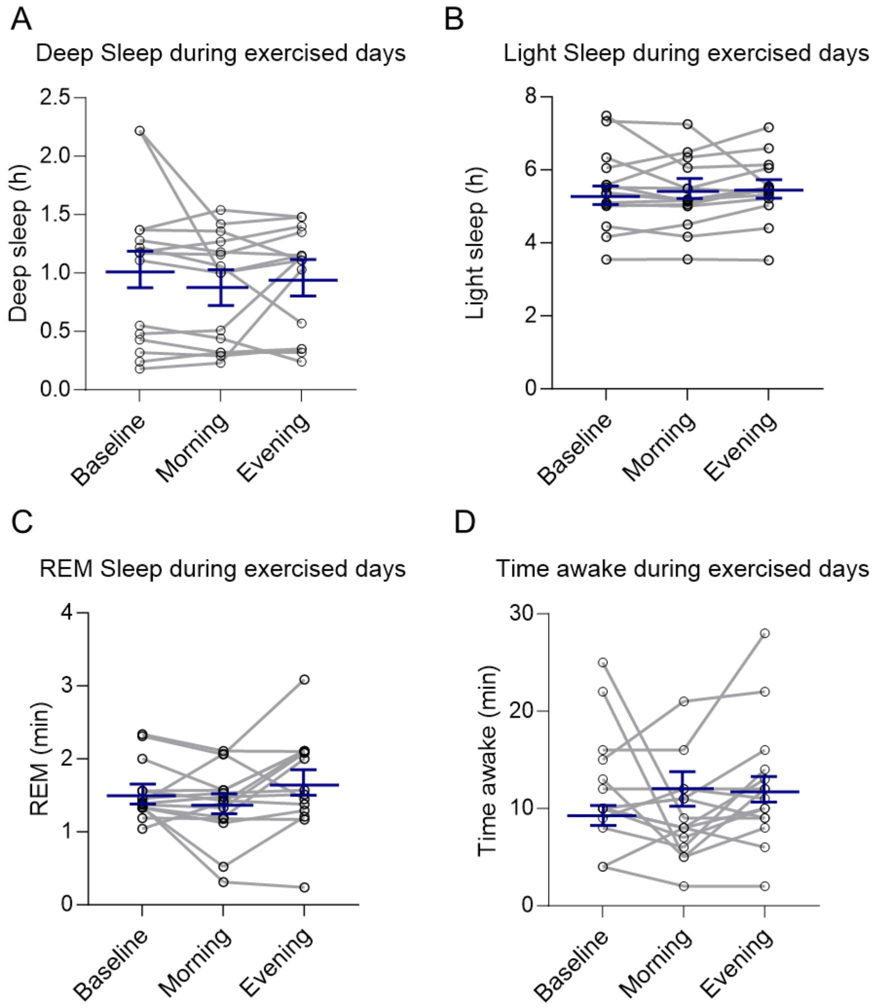


**Supplementary Figure 3**. Sleep architecture in the baseline, and morning and evening exercise periods of the trial. The values are shown (A) Deep sleep (n=15), (B) Light sleep (n=15), (C) REM sleep (n=15) and (D) Time awake (n=15), blue lines are mean±SEM. Data were analysed using one-way ANOVA followed up by Holm-Šídák's multiple comparisons test.

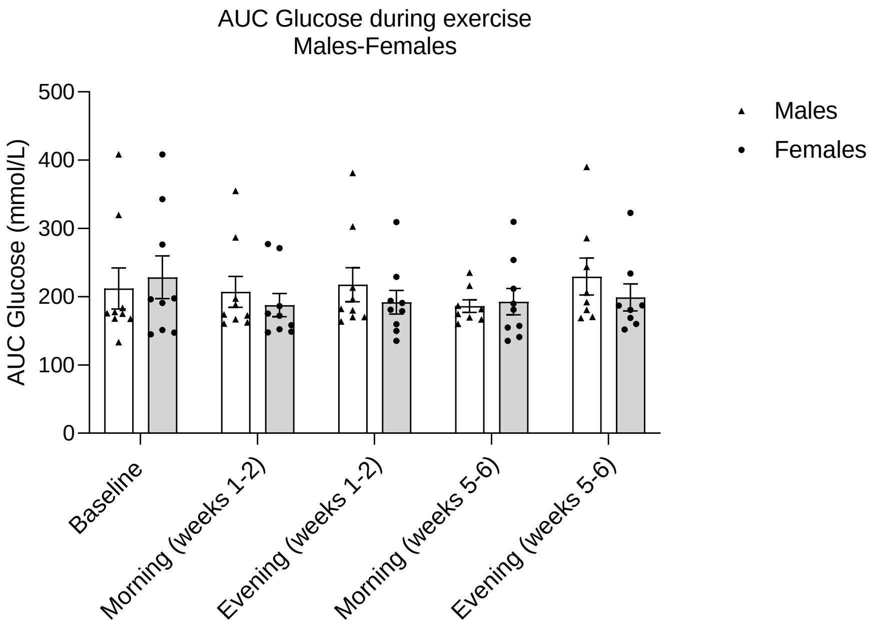


**Supplementary figure 4**. Mean area under the curve (AUC) glucose concentrations over 24-hours hourly during baseline, and morning and evening exercise split by sex. Glucose levels in Males weeks 1-2 (n=9), weeks 5-6 (n=8); Females weeks 1-2 (n=9), weeks 5-6 (n=8). Values are mean±SEM. Data were analysed using two-way mixed-model ANOVA followed up by Holm-Šídák's multiple comparisons test.

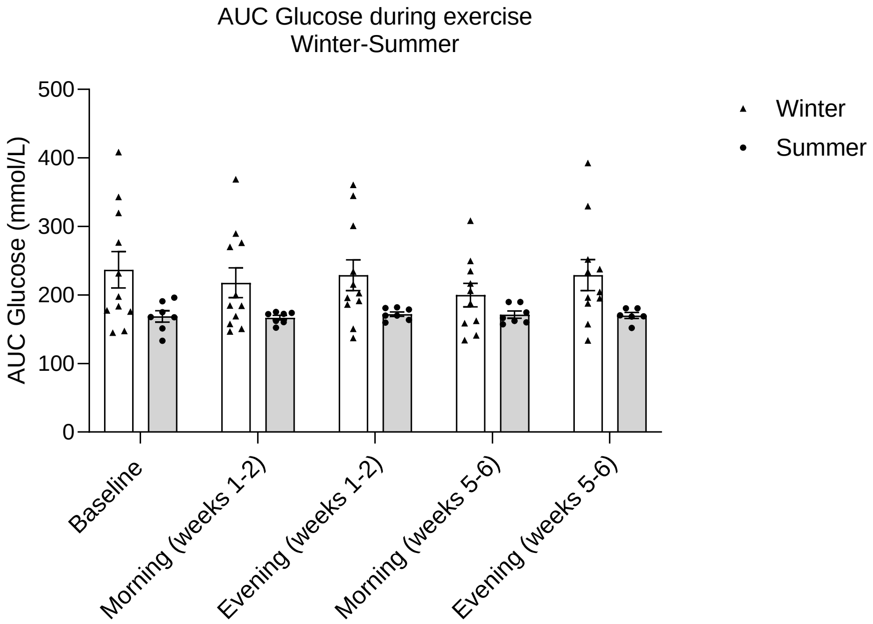


**Supplementary Figure 5**. Mean area under the curve (AUC) values over 24-hours hourly in baseline, and morning and evening exercise periods split by season. Winter weeks 1-2 (n=11), weeks 5-6 (n=10). Summer weeks 1-2 (n=7), weeks 5-6 (n=6). Values are mean±SEM. Data were analysed using two-way mixed-model ANOVA followed up by Holm-Šídák's multiple comparisons test.

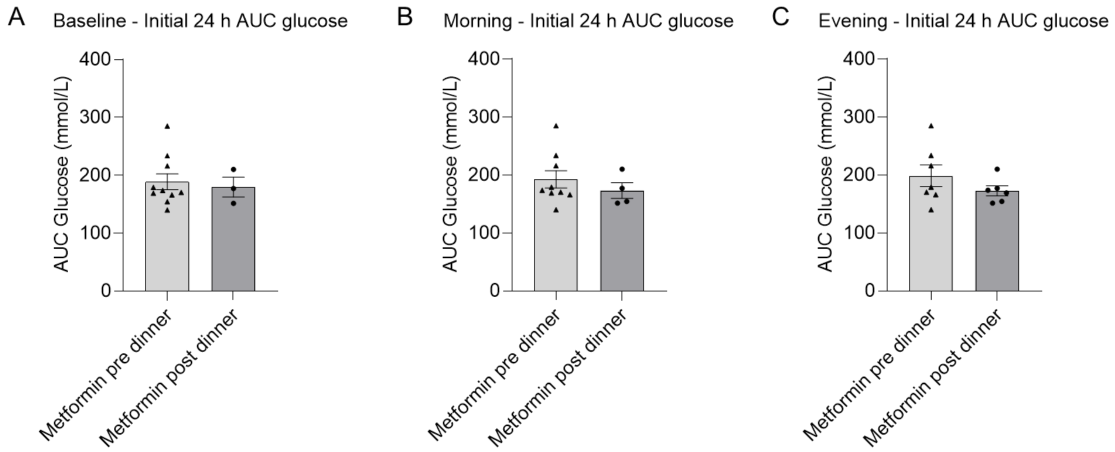


**Supplementary Figure 6.** Meal and metformin timing. Mean area under the curve (AUC) glucose values for the first 24 hours of exercise day comparing Metformin pre and post dinner. (A) Baseline, metformin pre-dinner (n=10), metformin post-dinner (n=3); (B) Morning exercise, metformin pre-dinner (n=9), metformin post-dinner (n=4); (C) Evening exercise, metformin pre-dinner (n=7), metformin post-dinner (n=6). Values are mean±SEM. Data were analysed using unpaired t-test.
